# Associations between temperament profiles, eating habits and BMI-SDS development in early childhood

**DOI:** 10.64898/2025.12.03.25341535

**Authors:** Saija Tarro, Jetro J. Tuulari, Akie Yada, Denise Ollas-Skogster, Maryam Zarra-Nezhad, Laura Perasto, Linnea Karlsson, Hasse Karlsson, Riikka Korja, Saara Nolvi, Minna Lukkarinen

## Abstract

**Background:** Early temperament may shape children’s eating behaviours and growth, yet longitudinal evidence supporting these associations remains limited. We aimed to identify latent temperament profiles from 6 months to 5 years and examine their associations with eating behaviours and longitudinal changes in BMI-SDS.

**Methods:** Data were drawn from the FinnBrain Birth Cohort Study, a population based longitudinal study in Southwest Finland (n=2,286). Child temperament was assessed at 6 and 12 months, and at 2, 4, and 5 years using validated questionnaires. Eating behaviour was reported at age 2, and BMI-SDS was measured at ages 2 and 5. Latent profile analysis was conducted on repeated temperament measures. Associations with outcomes were examined using the BCH method.

**Results:** Three temperament profiles emerged: *Dysregulated group* (38%; low self-regulation, low to average positive reactivity, low negative reactivity), *High self-regulation group* (37%: high self-regulation, low negative reactivity, high to average positive reactivity), *High negative reactivity group* (24%: high negative reactivity, moderate positive reactivity, average self-regulation). The *high self-regulation group* showed more favorable eating behaviours (less snacking and fussiness, more willingness to taste new foods) compared to the other groups. *High negative reactivity group* showed greater decrease in BMI than *Dysregulated group*. Other BMI change differences were non-significant.

**Conclusions:** Distinct temperament profiles were linked to early eating behaviours and, to a lesser extent, BMI development. Understanding these early temperament-based patterns may help identify children at risk for less favourable eating patterns and inform tailored early interventions to promote healthy eating and growth.

## 1. Background

Pediatric obesity is a major public health concern globally and associated with a range of obesity-related diseases and psychosocial burden (Rankin et al., 2016; Swinburn et al., 2011). In Europe, almost one out of every four preschool-aged children lives with overweight or obesity (Garrido-Miguel et al., 2019). Children living with obesity are more likely to become adults living with obesity (Simmonds et al., 2016). Thus, there is a need to understand early childhood factors at the child level that influence eating behaviours and the development of overweight, as these may increase risk in obesogenic environments (Harrison et al., 2011).

Temperament refers to biologically based individual differences in reactivity and self-regulation that are biologically based and relatively stable across contexts, i.e. trait-like (Rothbart, 2007, 2011; Rothbart & Bates, 2006). Reactivity is defined as the ease of motor and affective arousal and is described in two dimensions: positive and negative emotional reactivity (Putnam et al., 2006). Positive reactivity or affectivity, also referred to as surgency, includes the tendency to experience positive emotions, but also impulsivity and high intensity pleasure, approach behaviours, perceptual sensitivity, and low levels of shyness (Gartstein & Rothbart, 2003). In contrast, negative reactivity or affectivity is characterised by tendency experience and express negative emotions, such as anger, frustration, fear and sadness (Shiner et al., 2012). Self-regulation, i.e. effortful control, modulates reactivity and is defined by the capacity to refrain from a desired behaviour and maintaining attention on a task and resisting distraction (Putnam et al., 2006).

Temperament has been suggested as an early childhood factor that may be influence obesogenic eating behaviours and contribute to the development of overweight in children (Anzman-Frasca et al., 2012; Bergmeier et al., 2013; Slining et al., 2009; Stifter & Moding, 2019). Previous studies suggest that greater levels of negative reactivity in early life may increase the risk of overweight, and greater self-regulation may be protective (Anzman-Frasca et al., 2012; Skogheim & Vollrath, 2015; Stifter & Moding, 2019). Better self-regulation during childhood have been associated with a lower body-mass index in adulthood (Schlam et al., 2013).

Similarly to temperament, eating behaviours are partly biologically based and evolve throughout early childhood (Russell et al., 2023). These behaviours can be understood as incorporating both state-like and trait-like aspects, where enduring trait tendencies may shape responses to situational states (Russell et al., 2023). Self-regulation may be linked to obesity through excessive food intake; children with lower self-regulation may find it more difficult to resist highly palatable foods (Anzman-Frasca et al., 2012). Negative reactivity has been linked to both emotional overeating and undereating, suggesting that affective reactivity may influence eating regulation in complex and context-dependent ways (Messerli-Bürgy et al., 2018). Further, negative reactivity, especially fearfulness and withdrawal, has been identified as the temperament dimension most relevant to food neophobia (Moding & Stifter, 2016; Pliner & Loewen, 1997). However, recent evidence indicates that associations between child temperament and eating behaviour are shaped by contextual factors, including parenting practices and the broader eating environment (Tauriello et al., 2023). Moreover, these links may vary across developmental stages, with different patterns emerging in infancy, toddlerhood, and later childhood (Tauriello et al., 2023). Thus, more longitudinal research is needed assessing the relationships between temperament, eating and weight development from infancy to later childhood.

Although temperament is a relatively stable construct across development, early reactivity and emerging self-regulatory skills develop into greater voluntary control (Rothbart & Bates, 2006). Continuity has typically been investigated at the level of individual temperament dimensions (Carranza et al., 2013). However, these dimensions rarely operate in isolation but co-occur in meaningful patterns within individuals. A person-centered approach enables the examination of temperament continuity through profiles that capture combinations of dimensions and their stability or change over time (Carranza et al., 2013; Gartstein et al., 2017). To date, there is a lack of studies using person-centered analyses with longitudinal data spanning infancy, toddlerhood and the preschool years. Such longitudinal approaches are essential for understanding how temperament contributes to behavioural outcomes.

To address gaps in the current literature, we first aimed to identify latent temperament profiles from 6 months to 5 years of age. Second, we examined whether these profiles are associated with children’s eating behaviour. Third, we investigated associations between temperament profiles and BMI-SDS development in childhood.

## 2. Methods

### 2.1. Study design and subjects

The present study is based on data from children participating in the FinnBrain Birth Cohort Study. The study design has been described in detail previously (Karlsson et al., 2018). The study subjects were recruited during the first trimester at maternal welfare clinics by personal contact with a research nurse in the former South-West Finland Hospital District during 2012 to 2015. The FinnBrain study was approved by the Ethics Committee of Hospital District of Southwest Finland (57/180/2011).

For the first study question, all families who answered to temperament questionnaires at 6 months, 1 year, 2 years, 4 or 5 years of age were included in the sample. In addition, to the second and third study question all families who answered to eating habits questionnaire at 2 years of age and had data of child BMI, were included in the sample. Demographic characteristics of the study sample are presented in Table 1.

**Table 1.** Demographic characteristics of the study sample.

|  | **N (%)** | **Mean (SD)** | **Range** |
| --- | --- | --- | --- |
| Child sex, girl | 2 286 (46.8) |  |  |
| Mother BMI | 2 215 | 24.6 (4.7) | 16.6 - 60.6 |
| Gestational age | 2 286 | 39.8 (1.7) | 25.7 - 43.4 |
| Family monthly net income | 1 450 | 4 634 (3 863) | 500 - 60 000 |
| Family education | 2 242 |  |  |
| University degree | 1 014 (45.2) |  |  |
| Applied university degree | 662 (29.5) |  |  |
| Basic education | 566 (25.2) |  |  |
| Maternal depression | 1 774 | 4.9 (4.1) | 0 – 27 |
| Maternal anxiety | 1 775 | 3.7 (4.3) | 0 – 40 |

### 2.2. Measures

#### 2.2.1. Temperament

Mothers reported on children’s temperament at 6 and 12 months, and at 2, 4, and 5 years of age. At 6 and 12 months, the Infant Behaviour Questionnaire – Revised Short Form (IBQ-R SF, Gartstein & Rothbart, 2003; Putnam et al., 2014) was administered; at 2 years, the Early Childhood Behaviour Questionnaire (ECBQ SF, Putnam et al., 2006) and at 4 and 5 years, the Child Behaviour Questionnaire – very short form (CBQ VSF Rothbart et al., 2001). Parents were asked to assess their child’s behavior across a range of everyday situations during the previous week, using a 7-point scale ranging from “never” to “always”. From these questions, scores on three broad temperament dimensions were derived: positive reactivity, negative reactivity, and self-regulation.

The IBQ-R SF has 91 items containing 14 subscales forming three dimensions: approach, activity level, vocal reactivity, high intensity pleasure, smiling and laughter, and perceptual sensitivity form the dimension positive reactivity (Cronbach’s Alpha α for 6- and 12-month measures = 0.88 and 0.87, respectively), sadness, distress to limitations, fear and falling reactivity form the factor negative reactivity (α = 0.85 for both) and low intensity pleasure, soothability, cuddliness and duration of orienting form the factor self-regulation (α = 0.83 for both).

The ECBQ SF contains 107 items containing 18 subscales: approach, activity level, high intensity pleasure, impulsivity and sociability form the dimension positive reactivity (α = 0.82), discomfort, fear, frustration, motor activation, perceptual sensitivity, sadness, shyness and soothability for the dimension negative reactivity (α = 0.87) and attentional focusing, attentional shifting, cuddliness, inhibitory control and low intensity pleasure for the dimension self-regulation (α = 0.86).

The CBQ VSF contains 36 items containing 13 subscales: activity level, high intensity pleasure, impulsivity and shyness form the dimension positive reactivity (α for 4- and 5-year measures = 0.80 and 0.81, respectively), anger, discomfort, soothability, fear and sadness form the dimension negative reactivity (α = 0.74 and 0.77) and attentional focusing, inhibitory control, low intensity pleasure and perceptual sensitivity for the dimension self-regulation (α = 0.78 for both).

#### 2.2.2. Child eating behaviours

Children’s eating behaviours were assessed at age 2 using a parent-reported questionnaire consisting of 20 items on child’s dietary and oral hygiene habits, with an emphasis on behaviours relevant to oral health. The questionnaire did not capture all eating behaviours typically associated with obesity risk (e.g., fast-food consumption). Mothers reported the frequency of specific behaviours during the past month using a 7-point response scale (1=3-4 times per day, 2=twice per day, 3=once per day, 4=2-3 times per week, 5=once per week, 6=twice per month, 7=rarely or never). For this study, a subset of items was selected to reflect eating behaviour patterns related to appetite regulation, food approach and food avoidance. Snacking (eating outside regular meals) and consumption of sweet treats were used as indicators of impulsive or hedonic eating; food fussiness as indicator of food avoidance; and willingness to try new foods as a proxy for openness to new tastes and lower neophobia. Willingness to try new foods was reverse coded (7=1 … 1=7) so that lower scores consistently indicated more problematic or avoidant eating behaviours, thereby aligning all variables on the same direction.

#### 2.2.3. BMI

At the child ages of two and five years, BMI standard deviation scores (BMI-SDS) were calculated. Based on BMI-SDS, the Finnish weight classifications were used to combine weight classes into the dichotomy as follows: children with overweight, obesity or severe obesity vs. normal and underweight children (Cole et al., 2000; Saari et al., 2011). Furthermore, the longitudinal weight change was studied in two ways: 1) as the proportional weight between two and five years (normal weight vs. overweight during entire period) and 2) as the change of BMI-SDS between two and five years (ΔBMI-SDS) illustrating if child loses or gains weight (- or +).

### 2.3. Covariates

To control for other variables that have been observed to relate to eating habits and BMI, we chose to include infant biological sex assigned at birth, gestational age, maternal BMI, family education and income, and maternal psychological distress as covariates in the analyses. Data regarding sex, gestational age and maternal pre-pregnancy BMI were obtained from the Wellbeing Services County of Southwest Finland (VARHA) records. Families self-reported their monthly net income (EUR) and educational level in a questionnaire package at 5 years of age. Education was used as continuous variable in the analysis. The data on educational level was coded categorically as follows: 1□= basic education (high school-level or vocational training), 2□=□applied university degree or 3□=□university degree. Maternal psychological distress was chosen considering it is of particular interest in the FinnBrain cohort study design (Karlsson et al., 2018) and as its potential influence on maternal reports of child temperament traits (Durbin & Wilson, 2012). Maternal psychological distress was assessed at 2 and 5 years postpartum with maternal self-reports of postnatal depressive symptoms using the Edinburg Postnatal Depression Scale (EPDS, Cox et al., 1987; Cronbach’s Alpha α 0.85-0.86) and anxiety symptoms using the anxiety subscale from the Symptom Checklist -90, (SCL-90, Derogatis et al., 1973; Cronbach’s Alpha α 0.84-0.86). For present study, mean scores across the two time points were calculated.

### 2.4. Statistical analysis

Data were analyzed using R version 4.3.1 2024.09.1. (R Core Team 2024) and Mplus 8.8 software with the Maximum Likelihood estimation and robust standard errors (MLR) estimator (Muthén, L.K. and Muthén, 1998-2017). Full Information Maximum Likelihood (FIML) was used to handle missing data because it is considered one of the most robust approaches for parameter estimation in the presence of missing values because it uses all available information in the dataset without imputing missing values (Enders & Bandalos, 2001). R was used for descriptive data analyses and zero-order correlation analysis (Spearman’s rho correlation). In the study p-values below 0.05 were considered significant.

Latent profile analysis (LPA) was conducted in Mplus to identify subgroups of children based on repeated measures of temperament. Child temperament variables were standardized z-scores prior to analysis to allow comparison across scales and time points. The number of latent profiles was established by increasing the number of subgroups in the LPA models and comparing fit indexes of the outputs with an increasing number of subgroups. Decision about the optimal number of groups was based on AIC, BIC, the posterior probability (i.e., the probability of an individual belonging to a group) for each latent group, and entropy rate indexing classification accuracy.

In the second step, the Bolck-Croon-Hagenaars (BCH; Bolck et al., 2004) approach was applied to examine the mean differences in children’s eating behaviours (snacking, sweet treats consumption, food fussiness, and willingness to try new foods) and BMI change between ages 2 and 5 years across the latent profiles. Child and family background variables were included as covariates (child sex, maternal BMI, family income, parental education, and maternal psychological distress) in the analysis. Differences between profiles in BMI and eating behaviours were tested using Wald tests of parameter constraints within the BCH approach. When overall Wald tests were significant, pairwise comparisons were examined to identify which profiles differed from each other. The model can be expressed as: Eating behaviour or BMI change∼Latent profiles+Sex+Maternal BMI+Income+Education+ EPDS+SCL

The BCH approach uses a two-step method where latent classes are first estimated and individuals are assigned class-specific weights that account for classification uncertainty. In the second step, these weights are used to relate classes to distal outcomes or covariates without shifting the latent class solution (Bakk & Kuha, 2021). The BCH method is recommended for examining associations between identified profiles and distal outcomes because it accounts for measurement error of the latent profiles and the unequal variances among the variables by using posterior probabilities as weighting to account for individual differences (Nylund□Gibson et al., 2019). The BCH method does not assume normality of the outcomes.

## 3. Results

Descriptive statistics regarding the measures used in the study are displayed in Table 2.

**Table 2.** Descriptive statistics for the measures used in the study.

| Measure (theoretical range) | N | Mean (SD) | Range |
| --- | --- | --- | --- |
| IBQ-R 6 months (1-7) |  |  |  |
| Positive reactivity | 1927 | 4.8 (.7) | 2.4 - 6.6 |
| Negative reactivity | 1927 | 3.0 (.8) | 1.2 - 6.5 |
| Self-regulation | 1923 | 5.3 (.7) | 3.0 - 7.0 |
| IBQ-R 12 months (1-7) |  |  |  |
| Positive reactivity | 1636 | 5.1 (.6) | 3.3 - 6.5 |
| Negative reactivity | 1631 | 3.3 (.7) | 1.2 - 5.5 |
| Self-regulation | 1636 | 5.0 (.7) | 2.4 - 7.0 |
| ECBQ 2 years (1-7) |  |  |  |
| Positive reactivity | 1383 | 5.1 (.6) | 2.9 - 7.0 |
| Negative reactivity | 1384 | 2.9 (.6) | 1.7 - 5.7 |
| Self-regulation | 1391 | 5.0 (.6) | 3.0 - 6.6 |
| CBQ 4 years (1-7) |  |  |  |
| Positive reactivity | 1116 | 4.7 (.9) | 1.4 - 7.0 |
| Negative reactivity | 1117 | 3.5 (.8) | 1.4 - 6.4 |
| Self-regulation | 1117 | 5.3 (.8) | 2.1 - 7.0 |
| CBQ 5 years (1-7) |  |  |  |
| Positive reactivity | 1414 | 4.7 (.9) | 1.8 - 6.9 |
| Negative reactivity | 1417 | 3.6 (.9) | 1.0 - 6.4 |
| Self-regulation | 1423 | 5.5 (.8) | 2.4 - 7.0 |
| Eating behaviour (1-7) |  |  |  |
| Snacking | 1422 | 4.4 (1.4) | 1.0 – 7.0 |
| Consumption of sweet treats | 1420 | 4.8 (1.1) | 1.0 – 7.0 |
| Food Fussiness | 1425 | 4.1 (1.7) | 1.0 – 7.0 |
| Willingness to try new foods | 1417 | 4.0 (1.5) | 1.0 – 7.0 |
| Child BMI change between 2 - 5 years | 709 | -0.1 (.8) | -2.1 – 2.9 |
| Child BMI SDS at 2 years | 776 | 0.14 (1.1) | -4.2 – 3.2 |
| Child BMI SDS at 5 years | 733 | 0.06 (1.1) | -4.4 – 3.7 |

### 3.1. Zero-order correlations

Zero-order correlations among study continuous variables are shown in Figure 1. Maternal BMI correlated positively with child BMI-SDS at 2 and 5 years (Spearman’s ρ = 0.15 and 0.28, respectively), and with longitudinal BMI-SDS change (ρ = 0.17). In addition, maternal BMI showed modest associations with child snacking (ρ = -0.07). Child BMI-SDS at 2 and 5 years correlated positively with fussiness (ρ = 0.14 and 0.10, respectively). Family income correlated with maternal depression and anxiety (ρ = -0.15 and -0.12, respectively), as well as with child snacking (ρ = 0.09). Family education correlated with child snacking (ρ = 0.15). Maternal depressive and anxiety symptoms were correlated with child snacking (ρ = -0.14 and -0.11, respectively), and with fussiness (ρ = -0.15 and -0.13, respectively).

**Figure 1.**
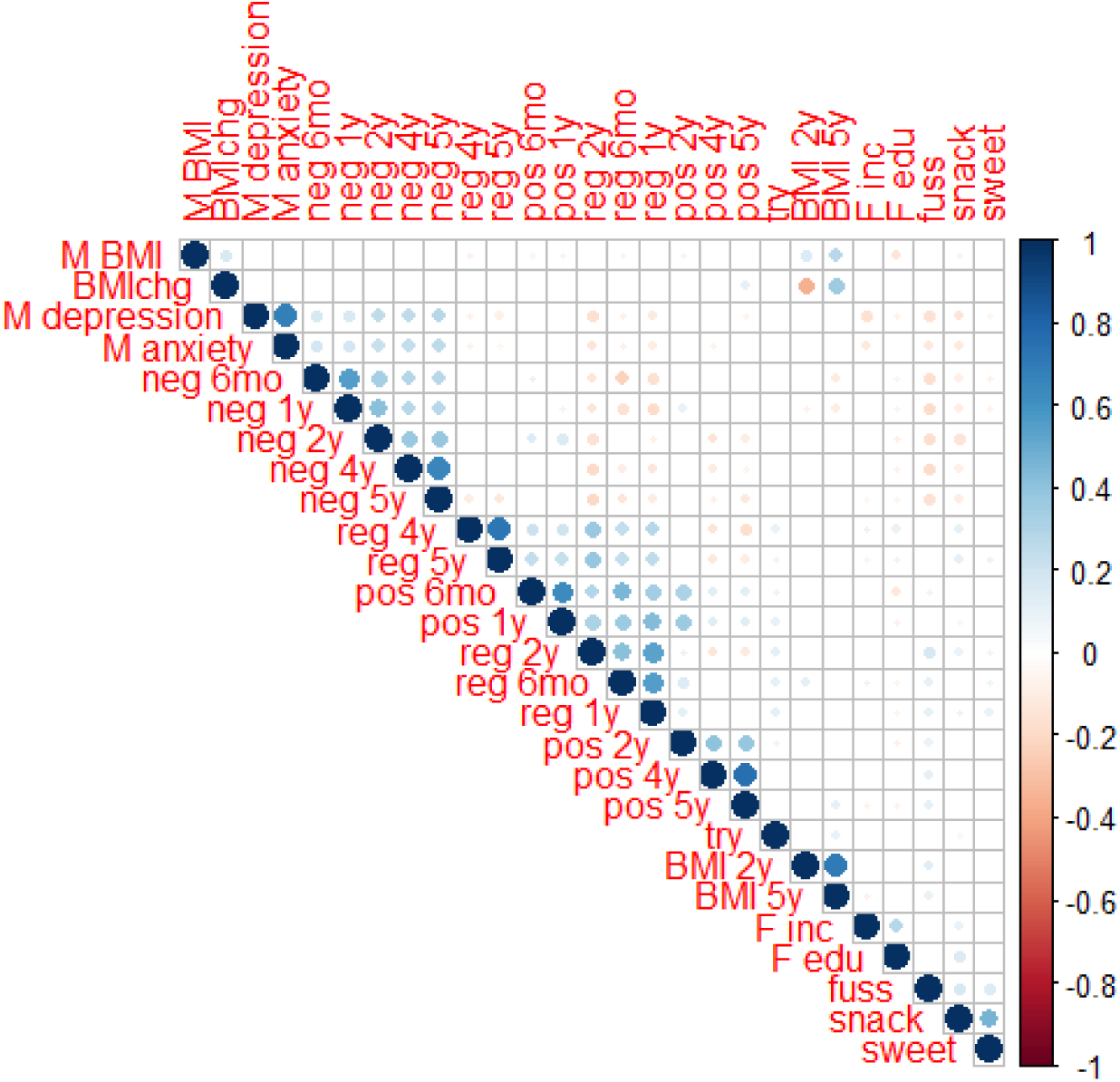
Zero-order correlations between measures in the study. Note. M = Mother, F = Family, C = Child, BMI = Body-mass index, gwk = Gestational age, inc = income, edu = education, pos = positive emotionality, neg = negative emotionality, reg = self-regulation, BMI chg = BMI change between 2 and 5 years of age, snack = snacking, sweets = consumption of sweet treats, fuss = food fussiness, try = willingness to try new foods. Only significant correlations (p < .05) are represented by a circle.

### 3.2 Latent Profile Solution

A latent profile analysis (LPA) based on child temperament dimensions (positive reactivity, negative reactivity, and self-regulation) identified a three-profile solution. The model demonstrated acceptable classification quality (entropy=0.65). The class proportions were as follows: group 1 (38%), group 2 (37%), group 3 (24%). Information of model fits can be found in Table 3.

**Table 3.** Model fits of latent profile analysis with increasing number of classes (n = 2.286).

| Class | AIC | BIC | saBIC | Entropy | VLMR<br>p value | LMR-A<br>p value | BLRT<br>p value | Class proportion | Average latent<br>class<br>probabilities |
| --- | --- | --- | --- | --- | --- | --- | --- | --- | --- |
| 1 | 63704.253 | 63876.29 | 63780.975 | 1 | - | - | - | 1.00 | 1.00 |
| 2 | 61691.432 | 61955.222 | 61809.072 | .653 | .0000 | .0000 | .0000 | .52/.48 | .89/.89 |
| 3 | 60897.143 | 61252.685 | 61055.7 | .65 | .0043 | .0045 | .0000 | .38/.37/.24 | .85/.85/.80 |
| 4 | 60363.592 | 60810.887 | 60563.067 | .634 | .4154 | .4177 | .0000 | .23/.34/.25/.18 | .77/.83/.77/.77 |
| 5 | 59935.725 | 60474.774 | 60176.119 | .638 | .2032 | .2042 | .0000 | .13/.26/.17/.22/.22 | .78/.74/.76/.80/.<br>76 |
| 6 | 59665.014 | 60295.816 | 59946.326 | .648 | .5582 | .5593 | .0000 | .12/.12/.28/.19/.14/<br>.15 | .81/.78/.72/.74/<br>.74/.77 |

Note. AIC = Akaike information criterion, BIC = Bayesian information criterion, saBIC = Sample adjusted BIC, VLMR = Vuong-Lo-Mendell-Rubin Likelihood Ratio Test, LMR-A = Lo-Mendell-Rubin Adjusted Likelihood Ratio Test, BLRT = Bootstrapped likelihood ratio test.

Group 1, *Dysregulated*, was characterized by low levels of self-regulation and negative reactivity combined with low to average positive reactivity (Figure 2). Group 2, *High self-regulation*, displayed high self-regulation, low negative reactivity and high to average positive reactivity, whereas group 3, *High negative reactivity*, was marked by high levels of negative reactivity, moderate positive reactivity, and average levels of self-regulation. Positive reactivity showed partial stability across childhood, with *High self-regulation group* maintaining consistently high levels, *Dysregulated group* remaining lower, and *High negative reactivity group* declining from initially high levels toward lower reactivity over time. In contrast, negative reactivity and self-regulation demonstrated notable stability, as the profiles maintained similar relative positions across all measurement points.

**Figure 2.**
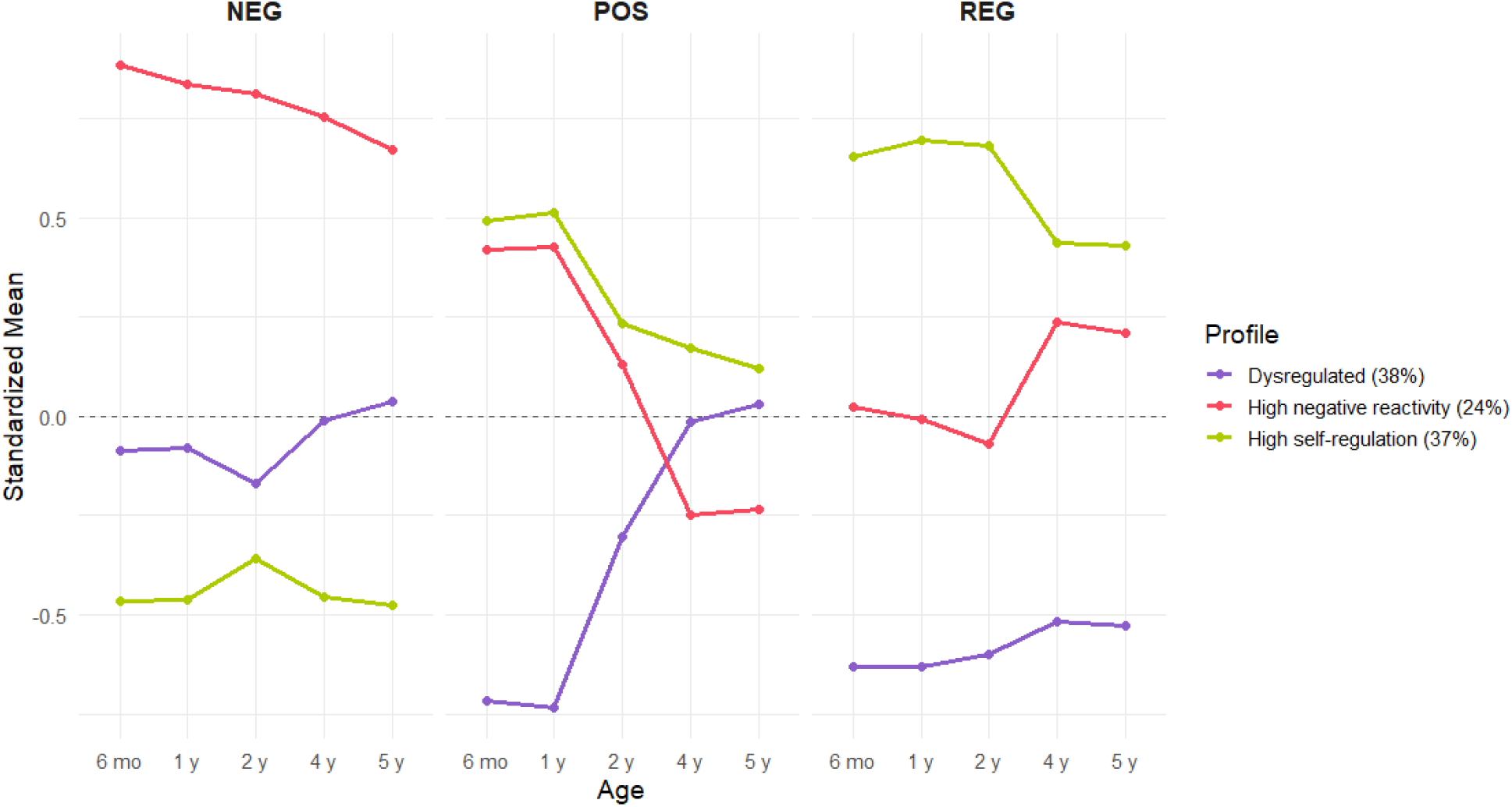
Standardized means of latent temperament profiles (Dysregulated, High self-regulation, High negative reactivity) in three dimensions (NEG = negative reactivity, POS = positive reactivity, and REG = Self-regulation) across ages 6 months, 1, 2, 4, and 5 years. Profiles were identified using latent profile analysis (LPA) and are displayed separately for each dimension. Percentages indicate profile prevalence in the sample.

Table 4 displays standardized means for three latent profiles (*Dysregulated, High self-regulation, High negative reactivity*) and their pairwise differences across dimensions (positive reactivity, negative reactivity, self-regulation) measured at multiple time points (6 months to 5 years). The largest differences are observed in negative reactivity dimension between groups *Dysregulated* and *High negative reactivity* and groups *High self-regulation* and *High negative reactivity*. Similarly, the majority of differences in self-regulation dimension are significant, indicating persistent and meaningful differences between profiles in regulatory capacity. In contrast, differences in positive reactivity are smaller, particularly between groups *High self-regulation* and *High negative reactivity*, indicating similar levels of positive reactivity across profiles.

**Table 4.** Means and pairwise differences between three temperament profiles.

|  | Group 1 | Group 2 | Group 3 | G1 vs G3 | G1 vs G2 | G2 vs G3 |
| --- | --- | --- | --- | --- | --- | --- |
| POS 6 mo | -0.718 | 0.493 | 0.421 | -1.139*** | -1.211*** | -0.072 |
| POS 1 y | -0.733 | 0.514 | 0.427 | -1.16*** | -1.247*** | -0.087 |
| POS 2 y | -0.302 | 0.234 | 0.129 | -0.431*** | -0.536*** | -0.105 |
| POS 4y | -0.015 | 0.171 | -0.248 | 0.233 | -0.186 | -0.419* |
| POS 5 y | 0.029 | 0.12 | -0.233 | 0.262 | -0.091 | -0.353* |
| NEG 6 mo | -0.086 | -0.467 | 0.884 | -0.97*** | 0.381* | 1.351*** |
| NEG 1 y | -0.081 | -0.463 | 0.838 | -0.919*** | 0.382* | 1.301*** |
| NEG 2 y | -0.169 | -0.36 | 0.813 | -0.982*** | 0.191 | 1.173*** |
| NEG 4y | -0.012 | -0.454 | 0.755 | -0.767*** | 0.442* | 1.209*** |
| NEG 5 y | 0.039 | -0.476 | 0.672 | -0.633*** | 0.515*** | 1.148*** |
| REG 6 mo | -0.629 | 0.655 | 0.022 | -0.651** | -1.284*** | -0.633*** |
| REG 1 y | -0.632 | 0.694 | -0.009 | -0.623*** | -1.326*** | -0.703*** |
| REG 2 y | -0.599 | 0.683 | -0.068 | -0.531* | -1.282*** | -0.751*** |
| REG 4 y | -0.517 | 0.436 | 0.237 | -0.754*** | -0.953*** | -0.199 |
| REG 5 y | -0.527 | 0.429 | 0.21 | -0.737*** | -0.956*** | -0.219* |

POS = positive reactivity, NEG = negative reactivity, and REG = Self-regulation Group 1 = Dysregulated, Group 2 = High self-regulation, Group 3 = High negative reactivity \* <0.05, \*\* <0.01, \*\*\* <0.001

### 3.3 Temperament profiles and eating behaviours

Adjusted mean comparisons indicated significant differences between the latent profiles on all four eating behaviour variables: snacking, sweet treat consumption, food fussiness, and willingness to try new foods (overall Wald tests: ps < 0.01). Pairwise comparisons, presented in Table 5, identify which profiles differ significantly on each behaviour. For snacking, children in *High self-regulation group* reported more favourable scores than both *Dysregulated* and *High negative reactivity group*, whereas *Dysregulated* and *High negative reactivity group*s did not differ significantly. For sweet treat consumption, *High self-regulation group* again showed the most favourable scores compared to other groups, with no difference between *Dysregulated* and *High negative reactivity group*s. Regarding fussiness, *High self-regulation group* displayed the lowest level of fussiness and was significantly more favourable than *Dysregulated* and *High negative reactivity group*s; *Dysregulated group* also scored more favourably than *High negative reactivity group*. Finally, for willingness to try new foods, both *High self-regulation* and *High negative reactivity group*s scored more favourably compared to *Dysregulated group*, whereas *High self-regulation* and *High negative reactivity group*s did not differ significantly.

**Table 5.** Comparisons among the temperament profiles for the child eating behaviour.

| Variable | Mean (SE) |  |  | Beta (SE) |  |  |
| --- | --- | --- | --- | --- | --- | --- |
|  | Group 1 | Group 2 | Group 3 | G1 vs G2 | G2 vs G3 | G1 vs G3 |
| Snacking | 5.19 (.40)<sup>1</sup> | 5.44 (.41) | 4.99 (.43) | -.25 (.11)<sup>*</sup> | .45 (.15)<sup>**</sup> | .20 (.14) |
| Sweet treats | 6.04 (.32) | 6.39 (.33) | 6.08 (.34) | -.35 (.09)<sup>***</sup> | .32 (.12)<sup>**</sup> | .04 (.12) |
| Fussiness | 5.22 (.50) | 5.79 (.52) | 4.82 (.53) | -.57 (.13)<sup>***</sup> | .97 (.17)<sup>***</sup> | .41 (.16)<sup>**</sup> |
| Trying new foods | 5.34 (.45) | 5.83 (.47) | 5.67 (.47) | -.49 (.12)<sup>***</sup> | .16 (.15) | .33 (.14)<sup>*</sup> |

<sup>\*</sup><0.05, <sup>\*\*</sup><0.01, <sup>\*\*\*</sup><0.001
<sup>1</sup> lower scores indicate more problematic or avoidant eating behaviours

### 3.4 Temperament profiles and BMI

Overall Wald tests indicated no significant differences in BMI across the three latent profiles (W = 4.31, df = 2, p = 0.12). Although the overall test was not significant, pairwise comparisons suggested that *High negative reactivity group* tended to have slightly greater decrease in BMI than *Dysregulated group*. Adjusted distal means for child BMI change between 2 and 5 years were -1.44 (SE = 0.41) for *Dysregulated group*, -1.51 (0.40) for *High self-regulation group*, and -1.67 (0.40) for *High negative reactivity group*. Girls showed a smaller decrease in BMI from 2 to 5 years compared to boys (β = 0.22, SE = 0.07, p = 0.001). Maternal BMI during pregnancy was positively associated with child BMI change (β = 0.04, SE = 0.01, p < 0.001), indicating that children of mothers with higher BMI showed smaller decrease in BMI compared to those of mothers with lower BMI. Other covariates were non-significant.

For BMI category, adjusted probabilities indicated that *Dysregulated group* had the highest likelihood of being in the reference category (0.72), *High self-regulation group* had the highest probability of constant obesity (0.18), and *High negative reactivity group* had the highest probability of early obesity (0.15). However, pairwise odds ratio comparisons between classes were not statistically significant.

## 4. Discussion

This study aimed to identify longitudinal temperament profiles from infancy to early childhood and to examine their associations with eating behaviour and BMI-SDS trajectories. We found three distinct temperament profiles across early childhood, and that distinct temperament profiles were associated with early eating behaviour and, to a lesser extent, with BMI development. These findings may highlight the potential for tailoring early interventions to temperament characteristics to promote healthy eating and prevent overweight.

Three temperament profiles emerged: *High self-regulation* (37%), characterized by low negative reactivity, high levels of self-regulation and high to average positive reactivity, *Dysregulated* (38%), marked by low levels of self-regulation, negative reactivity and positive reactivity, and *High negative reactivity* (24%), characterized by elevated negative reactivity, moderate positive reactivity and average self-regulation. Temperament characteristics similar to *High negative reactivity* and *Dysregulated* have previously been linked to heightened risks for emotional and behavioural problems (Abulizi et al., 2017; Bradley & Corwyn, 2008; Grady et al., 2012; Rothbart & Bates, 2006), suggesting that reduced regulatory capacity may underlie a range of adjustment difficulties, including less adaptive eating behaviours.

The present findings further suggest that children in the *High self-regulation* group displayed the most adaptive eating behaviour pattern, characterized by lower levels of snacking and sweet treat consumption, less fussiness, and greater willingness to try new foods. These behaviours likely reflect the combination of high self-regulation and low negative reactivity typical of this profile, which may facilitate regulation of appetite and openness to new experiences. In contrast, children in the *High negative reactivity* group consistently showed the least favourable pattern, marked by higher fussiness and less healthy eating behaviours. This may be explained by the heightened emotional reactivity and lower capacity to manage distress, which can lead to greater reliance on food for comfort and increased resistance to unfamiliar foods. Children in the *Dysregulated* group generally fell between these two extremes, showing less adaptive eating behaviours than the *High self-regulation group* but more favourable patterns than the *High negative reactivity* group. Their intermediate profile, characterized by moderate positive and negative reactivity combined with low self-regulation, may reflect challenges in maintaining emotional balance and behavioral control, which together can undermine consistent eating routines and openness to new foods. These differences across latent profiles align with previous research linking early temperament and self-regulation capacities with eating behaviours (Bergmeier et al., 2014a; Leung et al., 2014; Tauriello et al., 2023). Importantly, the present study extends the evidence by demonstrating these associations within a longitudinal, profile-based framework, highlighting how distinct combinations of temperament traits over time relate to eating behaviour patterns.

Across ages 2 to 5 years, BMI decreased on average in all temperament profiles, consistent with the normative developmental trajectory of BMI in early childhood (Wen et al., 2012). Although the overall test indicated no statistically significant differences between profiles, pairwise comparisons suggested a slightly greater decline in BMI-SDS in the *High negative reactivity* group compared to the *Dysregulated* group after adjusting for covariates. While these differences were not statistically significant, they may point to subtle variations in growth patterns that could be explored in future research. Previous studies have reported associations between difficult temperament (Niegel et al., 2007), surgency (Burton et al., 2011), distress to limitations (Darlington & Wright, 2006), and weight changes. However, such associations have typically been observed in infancy, and consistent evidence linking temperament with BMI beyond the first year of life remain limited. This developmental attenuation has led to the suggestion that temperament may exert a stronger influence on weight outcomes during infancy, when feeding interactions are primarily parent-driven, compared to later stages of development when child spend more time outside home in daycare and other settings (Bergmeier et al., 2014b; Tauriello et al., 2023). In later childhood, the relationship between temperament and BMI likely becomes indirect, operating through family dynamics and feeding practices. For example, child temperament has been shown to mediate the association between parental BMI and child BMI (Agras et al., 2004) and parental feeding behaviours may partially mediate the temperament-BMI link (Bergmeier et al., 2014b; Hittner et al., 2016). Further, low self-regulation has been linked with higher BMI only in boys who are living in a family high in household chaos (Riley et al., 2020). Parents of children with more “difficult” temperament might be more likely to use food as a soothing strategy (McMeekin et al., 2013). Restrictive feeding, in turn, has been identified as a risk factor for increased BMI gain in children, as it may disrupt children’s ability to self-regulate food intake (Jansen et al., 2012; Ventura & Birch, 2008). Together, these findings suggest that temperament may influence weight trajectories primarily through its impact on parent-child feeding interactions and the development of self-regulatory skills, rather than through direct physiological pathways.

A key strength of this study is the large sample size, which enhances statistical power and supports the generalizability of the findings. In addition, this study is part of a prospective birth cohort study that has a sample relatively representative of the Finnish population. Another important strength was that child temperament was assessed at multiple time points, allowing for a more robust and developmentally informed characterization of temperament profiles. Together, these features provide a strong basis for examining how early-emerging individual differences relate to subsequent eating behaviours and growth patterns. Several limitations should also be acknowledged. First, children’s eating behaviour was assessed only at the age of two using a limited number of questionnaire items, which may have constrained the scope and reliability of the construct captured and limited the ability to assess stability over time. Second, a notable amount of missing data for BMI outcomes likely reduced statistical power and precision of estimates. Third, while the latent profile approach enabled a holistic view of temperament across time, it did not capture the influence of finer-grained temperament facets or their potential interactions with specific eating behaviours and BMI.

Conceptually, our findings highlight the need for more refined theoretical frameworks linking temperament and eating behaviour. Just as previous work has advanced the conceptualization of temperament, comparable efforts are needed for nutrition behaviour (Lien et al., 2025). Future research should integrate biopsychosocial, neuropsychological, and physiological perspectives, while adopting refined definitions, phenotypic approaches, and multi-method assessments (Russell et al., 2023). Following these trajectories into later childhood and adolescence will clarify how early temperament-based differences in regulation and affectivity contribute to enduring patterns of diet quality and weight development. In this cohort, future analyses will examine how early temperament profiles relate to overall diet quality at ages five and nine and extend longitudinal modelling of temperament up to nine years of age. Importantly, interventions tailored to children’s temperament characteristics – for instance, supporting parents of children with more dysregulated temperaments in establishing consistent routines and responsive feeding practices – may help promote healthier eating trajectories and reduce later obesity risk.

## 5. Conclusions

This study identified three longitudinal temperament profiles in early childhood – *Dysregulated, High self-regulation, and High negative reactivity* – within the FinnBrain Birth Cohort. These profiles were differentially associated with children’s early eating behaviours, with the *High self-regulation* group showing the most adaptive patterns and the *High negative affectivity* group the least, and were only modestly related to BMI development. Establishing these temperament-based profiles provides a developmental framework for understanding how emerging temperament traits shape children’s eating behaviours and weight trajectories. The current findings highlight the importance of identifying targets for personalized interventions early, when BMI has not yet altered and eating habits can still be shaped. Supporting children based on their temperament profiles, while also considering family and environmental factors, could help prevent unhealthy eating habits and encourage healthy growth. As the cohort continues to be followed, extending this analysis into later childhood will enable a more comprehensive understanding of how early-emerging temperament traits may confer risk or protection in the development of healthy eating habits and weight trajectories.

## Declarations

### Funding sources

This work was supported by the Research council of Finland [grant numbers 308176 (LK), 264363 (HK), 253270 (HK), 308252 (RK), 308589 (LK), 358924 (MZ-N) and 358947 (MZ-N)], Finnish State Grants for Clinical Research (VTR – HK, LK), the Signe and Ane Gyllenberg Foundation (HK, LK), the Jane and Aatos Erkko Foundation (HK), the Stiftelsen Eschnerska Frilasarettet sr (HK), the Finnish Cultural Foundation (LK) and the Juho Vainio Foundation (ST). The funding bodies played no role in the study design, in the collection, analysis, and interpretation of the data; in the writing of the report; and in any restrictions regarding the submission of the report for publication.

## Acknowledgements

We thank the study families who took part in this study and the whole FinnBrain study research team for the data collection.

## AI use statement

An AI-based language model (ChatGPT) was used to assist with language editing. All AI-assisted outputs were carefully checked and verified by the authors.

## Author Contributions

All authors are responsible for the reported research. ST, JJT, RK, SN and ML contributed to the conceptualization and data collection of this study. ST, AY and LP contributed to the data analysis. Original draft was written by ST. LK and HK established the FinnBrain Birth Cohort and provided funding. All authors contributed to revision of the manuscript. All authors read and approved the final manuscript.

## Data availability statement

Finnish legislation on medical research and the ethics regulations specific to the FinnBrain cohort preclude open sharing of the original data sets. Inquiries on research collaboration and subsequent access to the data can be made to the Principal Investigator of the FinnBrain Cohort, Professor Linnea Karlsson.

## Abbreviations

BMI: Body Mass Index
CBQ: Child Behaviour Questionnaire
ECBQ: Early Childhood Behaviour Questionnaire
IBQ-R: Infant Behaviour Questionnaire-Revised
SDS: Standard Deviation Score

## Notes

### Competing Interest Statement

The authors have declared no competing interest.

### Author Declarations

The FinnBrain study was approved by the Ethics Committee of Hospital District of Southwest Finland (57/180/2011).

